# Unravelling habituation for COVID-19-related information: A panel data study in Japan

**DOI:** 10.1101/2022.08.15.22278703

**Authors:** Shinya Fukui

## Abstract

This study examines people’s habituation to COVID-19-related information over almost three years. Using publicly available data from 47 Japanese prefectures, we analyse how human mobility responded to COVID-19-related information, such as the number of COVID-19-infected cases and the declaration of a state of emergency (DSE), using an interactive effects model, which is a type of panel data regression. The results show that Japanese citizens were generally fearful and cautious in the first wave of an unknown infection; however, they gradually became habituated to similar infection information during subsequent waves. Nevertheless, the level of habituation decreased in response to different types of infections, such as new variants. By contrast, regarding the DSE, it is more plausible to consider that human mobility responds to varying requests rather than habituate them. We also find spatial spillovers of infection information on human mobility using a spatial weight matrix included in the regression model. The implementation of flexible human mobility control policies by closely monitoring human mobility can prevent excessive or insufficient mobility control requests. Such a flexible policy can efficiently suppress infection spread and prevent economic activity reduction more than necessary. These implications are useful for evidence-based policymaking during future pandemics.

## 1 Introduction

The COVID-19 pandemic has had significant socio-economic impacts at the global level as well as in Japan [1, 2]. Non-pharmaceutical interventions (NPIs), such as lockdowns, have been employed to suppress COVID-19 infections by restricting people’s outgoing behaviour [3-10]. Additionally, when infection spreads, people restrict their own outgoing behaviour, regardless whether NPIs are issued, to reduce risks [11–13]. For example, from March to May 2020, in the U.S., customer visits to the stores fell by 60%; 7% of which was explained by lockdowns, and the rest by own travel restrictions, owing to fear of infection [12].

As of July 2022, Japan experienced six waves of COVID-19 infection [14], and the government has issued four declarations of a state of emergency (DSE) as an NPI [15] between 2020 and 2021. In 2021, the government rolled out a rapid vaccination programme [16]. Additionally, the development of therapeutic agents has started worldwide [17], while much more has become known about post-infection effects [18]. However, new variants have emerged successively. Due to such a long-term pandemic, many individuals have grown tired of the COVID-19 pandemic, exhibiting the so-called ‘pandemic fatigue’ [19].

Throughout the COVID-19 pandemic, people have been exposed to two main pieces of information: the increase in the number of COVID-19 infections and NPIs, both leading them to feel fear and caution. As such, they changed their outgoing behaviours based on these two pieces of information [20–23]. Several studies thus explore the behavioural variations in response to COVID-19-related information over time. For example, Japanese citizens gradually reduced their stay-at-home response despite the repeated infection-increasing phase in 2020 [21, 24]. Similarly, using data from 124 countries, one study [19] observes a gradual decline in adherence to social distancing under the continued NPIs from March to December 2020. Other studies [25, 26] find that the extent of curtailment of going-out behaviours slowly decreased from the first to the fourth DSE. However, the human behavioural changes in response to COVID-19-related information, especially infection information, over almost three years, are still under-researched. This longer-term analysis is crucial for evidence-based policymaking for future pandemics.

Our study uses publicly available human mobility data and analyses how human behavioural responses to COVID-19-related information vary over the longer term. In other words, we explore whether people are habituated to the information. Habituation [27, 28] refers to the diminishing reaction to a repeated stimulus over time; in our case, the stimulus is represented by fear and caution [29]. For instance, a study [30] using individual questionnaires shows that COVID-19 anxiety became habituated over sixteen months. To investigate whether habituation has occurred, first, we examine the variation in human mobility in response to COVID-19 infection information over six waves; second, we re-examine the impact of multiple DSE on human behaviour. As an NPI in Japan, a DSE does not legally regulate behaviour. Accordingly, the decision to restrain oneself from going out is voluntary (see Details of DSE requests in Appendix A in the Supplementary Information).

Further, vaccination affects people’s fear and caution. Whether increased vaccination rates decrease people’s fear and caution of infection and promote outgoing behaviours [31] is also analysed. We also examine the spatial spillovers between prefectures, as changes in human mobility in one prefecture are influenced by information from other prefectures, using cross-prefecture travel (representing spatial interaction). Several studies have noted the importance of considering regional spatial interaction when investigating the impact of COVID-19 infection or NPIs [25, 32, 33]. As such, we incorporate a cross-term using a spatial weight matrix, which has not been used in previous related studies, to illustrate the spatial interactions between prefectures. Moreover, a single wave of infection comprises an increasing and decreasing phase. The difference between the two is that people are more likely to stay at home in the increasing phase but will gradually resume going out in the decreasing phase. Human mobility responses are considered heterogeneous across the two phases. We validate this conjecture for the first time by identifying the start, end, and peak of infection of different waves in each prefecture in Japan. We utilise the interactive effects model [34] in our regression analysis, which is to control for the unobservable factors which also affect human mobility.

Exploring how human mobility responded to (a) two pieces of information (i.e., repeated waves of infection or new variants and several DSEs), (b) the spatial spillovers of these two pieces, and (c) the different infection phases is important in considering mobility control policies over a long-term pandemic. While this study does not directly examine individual attitudes, it indirectly explores people’s habituation from COVID-19-related information, which arises due to decreased fear and caution, by assessing how human mobility responses changed during the pandemic.

## 2 Methods

### 2.1 Human mobility

The daily human mobility data for each prefecture used in this study are obtained from Google’s COVID-19 Community Mobility Reports [35], which are composed of six human mobility categories: *retail & recreation, grocery & pharmacy, parks, transit stations, workplaces*, and *residential*. From these, we select *retail & recreation* and *residential*. The data show the percentage change compared to a day-of-the-week baseline calculated based on median values for each day of the week for the five weeks from 3 January to 6 February 2020—just prior to the global outbreak of the COVID-19 pandemic. When avoiding unnecessary mobility, either in reducing the risk of infection or responding to the DSE by the government, people minimise their outings mainly to retail stores (not grocery stores and pharmacies, which are essential) and entertainment venues and also have a greater tendency to stay at home. In Google’s data, *residential* indicates time spent at home.

### 2.2 Regression model

Our analysis uses Bai’s ‘interactive effects model’ [34]. Suppose that we have a panel data set in which {(*y*_*it*_, *X*_*it*_)}, *i* = 1, 2, ⋯, *N, t* = 1, 2, ⋯, *T*; then, the interactive effects model is expressed as follows:

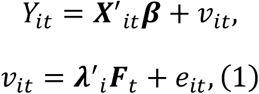

where *Y*_*it*_ is a dependent variable, ***X***_*it*_ is a *p* × 1 vector of explanatory variables such that *p* is the number of explanatory variables, and ***β*** is a *p* × 1 vector of the parameters to be estimated. ***λ***_*i*_ is a *d* × 1 vector of factor loadings that differ for unit *i* and ***F***_*t*_ is a *d* × 1 vector of a common factor that varies over time *t*, where *d* is the dimension of factors, and *e*_*it*_ is the error term. An unobservable term *v*_*it*_ has factor structure ***λ***^*’*^_*i*_ ***F***_*t*_ and random part *e*_*it*_.

Applying the interactive effects model (Eq (1)), our estimation model becomes:

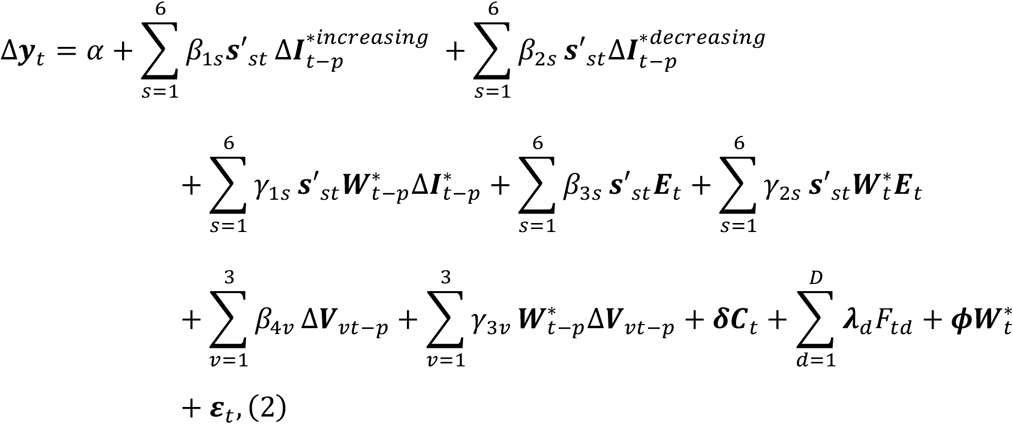

where **Δ*y***_*t*_ is an *N* × 1 (*N* = 47 prefectures) vector of the week-on-week difference in the percentage change from the baseline of human mobility or the residential time on day *t* (in percentage-points [pp]). Variable 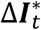 is an *N* × 1 vector of the week-on-week difference of new infections transformed by the inverse hyperbolic sine. This variable approximates the growth rate of new infections compared to the previous week. We separate 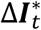 for the increasing 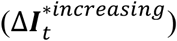 and decreasing 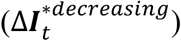 phases. ***s***_*st*_ is an *N* × 1 vector of the step dummies, which takes 1 with each corresponding wave, *s* = 1, 2, ⋯, 6. ***E***_*t*_ is an *N* × 1 vector of the DSE dummy that takes the value of 1 under a DSE and 0 otherwise. Since DSEs were only issued for the first, third, fourth, and fifth waves, the DSE dummies for the second and sixth waves are zero. Variable **Δ*V***_*vt*_ is an *N* × 1 vector of the week-on-week change in the vaccination rate per million persons. Index *v* denotes the number of vaccine doses (from the first to the third dose) administered.

Here, spatial weight matrices 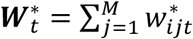 vary over time *t*, where *w*_*ij*_ is an element of the spatial weight matrix with row *i* (travel from) and column *j* (travel to), and diagonal element *w*_*ii*_ is 0, and represent the weekly time-series changes in the movement of people between prefectures. Therefore, cross-terms 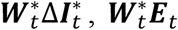, and 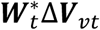 exhibit the impact of information such as the number of infections, the DSE, and the vaccination rate from other prefectures, respectively. The larger the spatial weight matrix elements are, the larger the impact on its own prefectures; that is, the more travel from the other prefecture, the more influenced by information from that prefecture. None of the studies that have explored human mobility regarding COVID-19 have considered these spatial interactions.

In the estimation, reverse causality from the dependent variable (**Δ*y***_*t*_, human mobility within one’s prefecture) to the spatial weight matrix (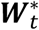, human travel between prefectures) is not a concern. As we use the movement from other prefectures to the relevant prefecture to construct the spatial weight matrix, human mobility in one’s own prefecture does not directly affect human travel from other prefectures to one’s prefecture.

Term ***C***_*t*_ is an *N* × ***K*** matrix of a control variable, where ***K*** is the number of control variables. ***F***_*td*_ represents the common factors of dimension *d*; ***λ***_*d*_ is an *N* × 1 vector where *d* = 1, 2, ⋯, *D* is the dimension of factors, representing factor loadings. This factor structure allows the proposed model to capture time-varying unobservable elements, such as people’s fear and caution of new variants emerging in some countries with different loadings over cross-sectional units, and can better describe these than ordinary two-way fixed effects. Meanwhile, 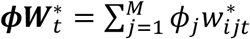 represents spatially weighted fixed effects [36, 37]. Finally, ***ε***_*t*_ is an i.i.d. (independent and identically distributed) random component vector. ***λ***_*d*_, ***F***_*td*_, and ***ε***_*t*_ are unobservable.

Parameters *α, β*_1*s*_, *β*_2*s*_, *β*_3*s*_, *β*_4*v*_, γ_1*s*_, γ_2*s*_, γ_3*v*_, ***δ*** (***K*** × 1 vector), and ***ϕ*** (*M* × 1 vector) are to be estimated, while the parameters of interest are *β*_1*s*_, *β*_2*s*_, *β*_3*s*_, *β*_4*v*_, γ_1*s*_, γ_2*s*_, and γ_3*v*_. The explanatory and control variables are further described in Appendix A in Supplementary Information. The regression model is described in detail in Appendix B in Supplementary Information.

### 2.3 Identifying COVID-19 waves

As the government made no official announcements regarding the beginning and end of COVID-19 infection waves, we independently determine the COVID-19 wave duration of each prefecture. Further details are provided in Appendix C in Supplementary Information.

## 3 Results

The empirical results for human mobility of retail and recreation are presented below.

### 3.1 Empirical approach

We use panel data from 47 prefectures over 850 days, from 22 February 2020 to 20 June 2022, for six infection waves. Lags (*p* in Eq (2)) are taken from 1 to 7 days for the week-on-week growth rate of new infections, week-on-week changes in vaccination rate, and the spatially weighted of these variables.

### 3.2 Human mobility and explanatory variables

Fig 1 displays time series plots of the variables for Tokyo and Osaka from among the 47 prefectures, two representative urban areas in Japan. The figure shows an inverse relationship between the number of infections and human mobility: as the number of infected people increases, human mobility tends to decrease, and vice versa. During the first infection wave, human mobility shows a sharp decline. Subsequently, even when the DSEs were issued, human mobility also declined sharply. In late 2021, human mobility recovered notably. However, human mobility decreased sharply, even without the DSE, when the number of infections surged around the beginning of 2022. The first and second vaccination doses were administered rapidly, without any gap between them. The above trends are common between Tokyo and Osaka.

**Fig 1.**
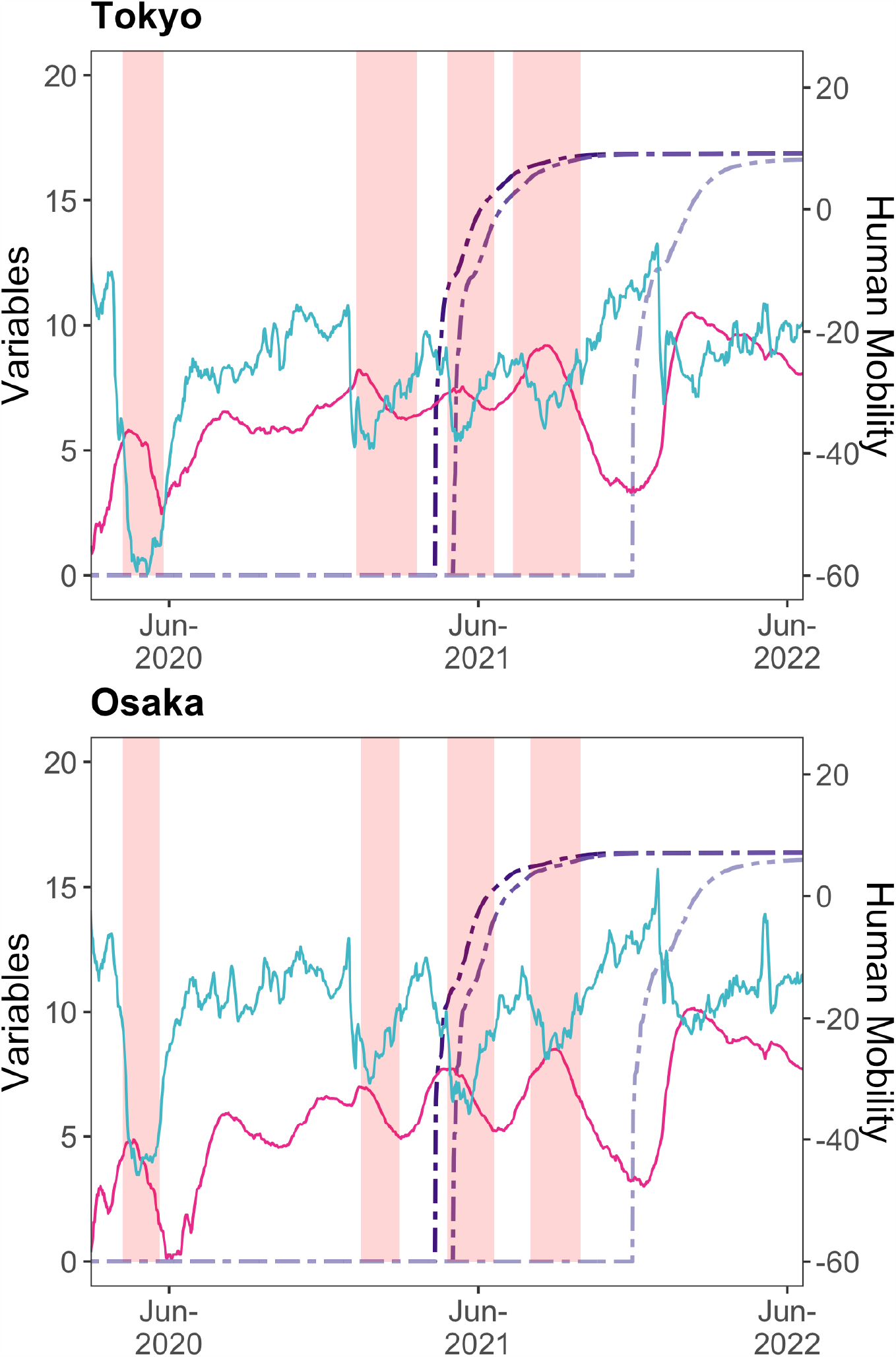
Time series plots of the variables for Tokyo and Osaka. On each chart, the blue-green-coloured line (on the right-hand axis) is the 7-day backward moving average using the geometric mean of human mobility in retail and recreation; the red-purple-coloured line is the IHS transformation of the 7-day backward moving average number of infected persons; the purple-coloured double-dashed lines are the IHS transformation of the cumulative number of people vaccinated with 1–3 doses, and the pink-shaded areas are the DSE periods. The data transformation employed here, such as the 7-day backward moving average, is only for visualisation purposes; we use other transformations in our estimation.

### 3.3 Retail and recreation human mobility response

#### 3.3.1 Infected cases in the increasing phase

As shown in Fig 2a, during the phase of increasing infections, the fear and caution of new infectious diseases substantially reduces human mobility in the first wave. A 1% week-on-week increase in the number of infected cases results in a decrease in human mobility (which is the percentage change from the baseline) by 1.09 pp week-on-week, at most (lag: 2, standard error [s.e.] = 0.13). The reduction in human mobility in the first wave weakened daily from 2-to 7-day lags. The 6-and 7-day lags have a wide confidence interval (CI) (95% CI for lag 7 = [-1.36 to -0.23]), indicating that human behaviours on these lag days varied widely among prefectures after receiving infection information.

**Fig 2.**
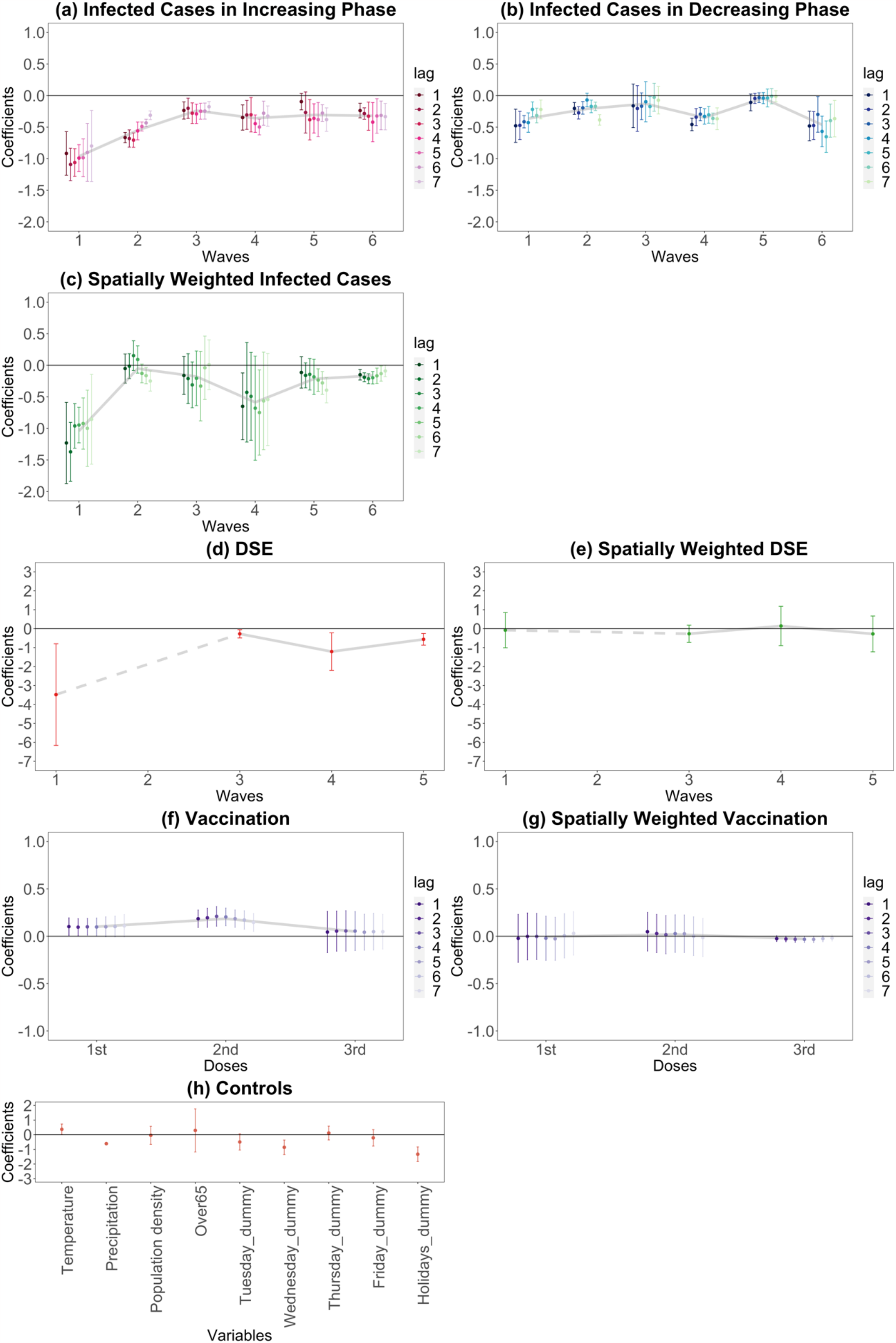
Retail and recreation human mobility responses to COVID-19-related information. On each chart, the points are estimated coefficients, and the bars indicate upper and lower 95% confidence intervals. The grey line traces the average coefficients of each lag day. There are six infection waves, but DSEs were only issued for the first, third, fourth, and fifth waves. We take a daily lag from 1 to 7 days for infected cases in the increasing phase, infected cases in the decreasing phase, spatially weighted infected cases, vaccination, and spatially weighted vaccination. We conduct the regression analysis seven times, from lags 1 to 7. We do not take a daily lag for the DSE, spatially weighted DSE, and control variables; these estimates are from the lag-1 regression.

The extent of reductions during the second wave is smaller than during the first, but it is still high with a maximum 0.71 pp decrease (lag: 3, s.e. = 0.06). The responses in the second wave also decreased daily, from 3-to 7-day lags. The third wave shows a more modest human mobility response than the first and second waves, indicating a tendency towards habituation, with a maximum 0.29 pp decrease (lag: 4, s.e. = 0.08). The response remained flat for each lag day.

Meanwhile, the human mobility response rose in the fourth wave; the maximum decrease is 0.50 pp (lag: 5, s.e. = 0.06) and the reduction is strengthened on lag days 4 and 5. In the fifth wave, although lag days 1 and 2 do not meet the 5% significance threshold, human mobility is largely reduced from the 3-to the 7-day lags with a maximum 0.38 pp decrease (lag: 3, s.e. = 0.16). The maximum decrease in the sixth wave is 0.42 pp (lag: 4, s.e. = 0.16) and the response gradually intensified from lag day 1 to 4, remaining significant until lag day 7, with similar reductions as in waves 4 and 5.

#### 3.3.2 Infected cases in the decreasing phase

During the phase of decreasing infection (the recovery phase), as presented in Fig 2b, if the negative range in the estimated value is large, it indicates a prominent week-on-week increase in human mobility (the percentage change from the baseline), responding to the decreasing infected cases. One explanation is that people may become less fearful and less cautious of infection in the decreasing phase. Another is that if human mobility decreases greatly during the increasing phase, there is a positive rebound during the recovery phase.

As a result, in the first wave, the negative range is relatively large. However, human mobility remains somewhat reduced, as the magnitudes of the estimates are smaller than those in the increasing phase. A 1% decrease in the number of infections will result in, at most, a 0.48 pp (lag: 1, s.e. = 0.13) increase in human mobility. In the second wave, the maximum is a 0.39 pp increase (lag: 7, s.e. = 0.04). Only lag 5 is significant at the 5% level for the third wave; however, the estimated coefficient is small.

By contrast, the decreasing phase of the fourth wave shows at most a 0.46 pp (lag: 1, s.e. = 0.05) increase in human mobility—a similar magnitude to that of the increasing phase (0.50 pp); it had recovered to the same extent that it decreased during the increasing phase during the fourth wave. However, in the fifth wave, the estimates are no longer significant and human mobility did not recover. Finally, the sixth wave shows the highest value of all waves, with a maximum increase of 0.65 pp (lag: 5, s.e. = 0.13).

#### 3.3.3 Spatially weighted infected cases

The increasing and decreasing phases are not separated for the spatially weighted infected cases (Fig 2c). We find that individual going-out behaviours dramatically changed in response to infection information from the other prefectures in the first wave, with the maximum (in absolute value) being 1.37 pp (lag: 2, s.e. = 0.24). The wider CI than the response for one’s own prefecture (Fig 2a,b) indicates that the impact from other prefectures varies greatly by prefecture. From the second to the third wave, the results are insignificant for most lags; the magnitudes are modest for those significant estimates. Conversely, in the fourth wave, lags 1 and 5 are significant at the 5% level; the maximum (in absolute value) is 0.75 pp (lag: 5, s.e. = 0.34); however, the CIs are wide. In the fifth wave, lags 5 to 7 are significant, with a maximum of 0.39 pp (in absolute value) (lag: 7, s.e. = 0.10). All lags are significant in the sixth wave, with the largest being 0.21 pp (in absolute value) (lag: 3, s.e. = 0.04). Therefore, the first and the fourth to the sixth waves affected going-out behaviours.

#### 3.3.4 DSE and spatially weighted DSE

A DSE was only issued for the first, third, fourth, and fifth waves (Fig 2d). The first DSE (in the first wave) greatly reduced human mobility. Although the CI is relatively large, we see a 3.48 pp week-on-week decrease (s.e. = 1.37) in the percentage change from the baseline human mobility. Although significant in the second DSE (third wave), the magnitude is low (0.27 pp decrease, s.e. = 0.11). In the third DSE (fourth wave), the magnitude is large, with a 1.21 pp decrease (s.e. = 0.51). The fourth DSE (fifth wave) lead to a slightly lower but still significant, with a 0.56 pp decrease (s.e. = 0.16). By contrast, the spatially weighted DSE is insignificant in any of the waves (Fig 2e).

#### 3.3.5 Vaccination and spatially weighted vaccination

Regarding vaccination, positive estimates are expected because as the vaccination rate increased from the previous week, the more secure people felt going out. As a result (Fig 2f), for a first vaccine dose, only some lags are significant, with each having a negligible impact. The effect is more apparent for the second than the first dose, which is significant for all lag days. A 1 pp week-on-week increase in the vaccination rate leads to up to 0.21 pp week-on-week increase in the percentage change from the baseline human mobility (lag: 3, s.e. = 0.05). However, none of the results is significant for the third dose. Spatially weighted vaccination is not significant for any doses (Fig 2g, only lag 3 in the third dose is significant, while the magnitude is negligible).

#### 3.3.6 Control variables

As for the control variables (Fig 2h), each day and holiday dummy are significant. Additionally, the precipitation coefficient is negative and significant, meaning increased precipitation reduced human mobility.

### 3.4 Robustness

To test robustness, we conduct the estimation via the polynomial degree 1 Almon lag model of retail and recreation mobility (Fig 3, detail of the Almon lag model is described in Appendix D in Supplementary Information). As shown in Fig 3, although vaccination is only significant at the second dose lag-4 (and although slightly different results for spatially weighted vaccinations), the results indicate a similar tendency to our main results. That is, regarding infected cases:

**Fig 3.**
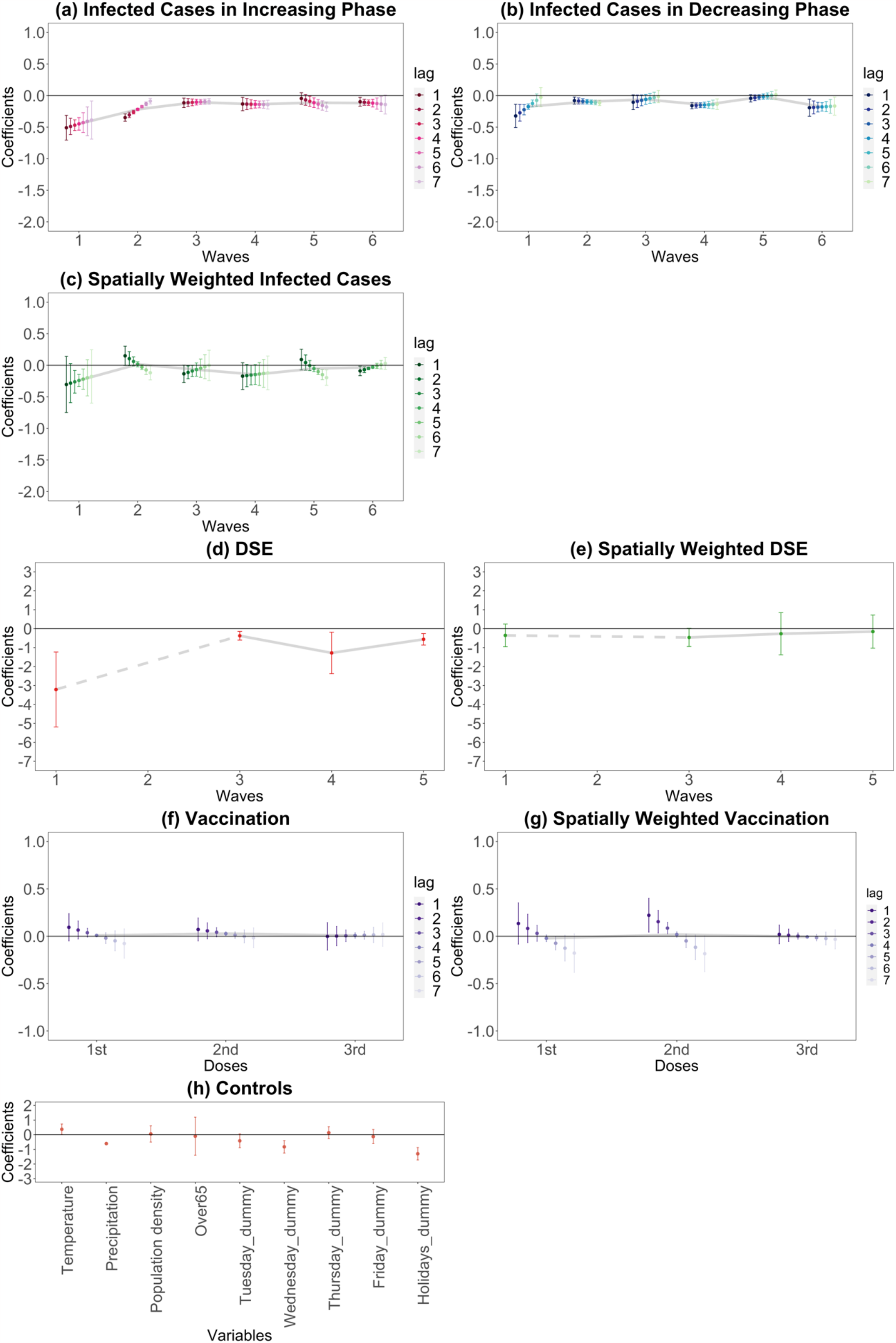
Retail and recreation human mobility responses to COVID-19-related information using the Almon lag model. On each chart, the points are estimated coefficients, and the bars indicate upper and lower 95% confidence intervals. The grey line traces the average coefficients of each lag day. There are six infection waves, but DSEs were only issued for the first, third, fourth, and fifth waves. We take a daily lag from 1 to 7 days for infected cases in the increasing phase, infected cases in the decreasing phase, spatially weighted infected cases, vaccination, and spatially weighted vaccination. We do not take a daily lag for the DSE, spatially weighted DSE, and control variables.

- Habituation of responses from the first to the third waves of the increasing phase
- Augmented responses in the fourth to the sixth waves
- Heterogeneity of responses in increasing and decreasing phases
- Lag day trend
- Spatial spillovers in the first, fourth, fifth, and sixth waves

Regarding DSEs and vaccination:

- Significant responses of DSEs and second-dose vaccinations in own prefectures

In addition, the results of the residential time, which we expect to reflect the opposite impact of that for retail and recreation, are shown in Figs. E1 and E2 in Appendix E in the Supplementary Information. These results confirm the robustness of our main results.

## 4 Discussion

Retail and recreation mobility responses during the increasing (exacerbating) infection phase suggest that, in the first wave, people largely feared and cautioned the increase in infection numbers. However, people gradually became accustomed to infections over the first three waves. This process can be described as ‘habituation’, that is, people becoming accustomed to similar infection information and gradually decreasing their fear and caution. However, smaller magnitudes of human mobility responses in the decreasing phase compared to the increasing one over the first three waves exhibit heterogeneous responses. This heterogeneity shows that people were cautious about the recovery of human mobility despite habituation trends in the increasing phase. Early in the pandemic, in 2020 and early 2021, the fear and caution of new infections were presumed to be strong. Incidentally, the reduction in human mobility during the first and second waves weakened daily, as people presumably tended to respond more strongly to the most recent information in the early stages.

By contrast, from the fourth to the sixth waves (from spring 2021 to June 2022), the Alpha, Delta, and Omicron variants began to dominate, respectively [38]. As a result, the responses of people during the increasing phase, which had declined until the third wave, began to strengthen again and the habituation level decreased. Each new variant exacerbated the speed of infectivity [39], which led to increased fear and caution. In addition, unlike in the first three waves, where the response weakened daily or remained flat, from the fourth to the sixth wave, the responses tended to intensify from lag day 3 to lag day 5 in each wave. Due to the emergence of new variants, people probably became much more reluctant to go out after receiving information about growing numbers of infections for successive days beyond their anticipation.

Interestingly, the decreasing phase of infection in wave four suggest a prominent recovery of human mobility, which again implies the heterogeneous response between the increasing and decreasing phases. This recovery is probably due to some positive events: the promotion of a rapid vaccination programme [16] and information regarding COVID-19 treatment and the post-infection situation [17, 18]. By contrast, in the fifth wave, the estimates are not significant in the decreasing phase, likely because the Delta variant is more transmissible and severe than the Alpha variant in the fourth wave [39], making people more fearful and cautious. Therefore, human mobility did not recover. In the sixth wave, the Omicron variant is prevalent and more transmissible than the previous ones, but less severe [39]. Therefore, people probably went out more when the number of infections decreased.

The effect of vaccination is apparent: increased second-dose vaccination rates led to the recovery of human mobility. At that time, the Japanese government released information on the efficacy of the second dose [40]. During the long-term COVID-19 pandemic, a rapid vaccination programme and its information dissemination help alleviate people’s concerns regarding COVID-19 infection and make them feel safe and secure. However, because the timing of the third vaccination varied greatly by person, human mobility does not show substantial recovery.

As for DSEs, it is more plausible to consider changes in the human mobility response as per varying requests rather than habituation (see Details on DSE in Appendix A in the Supplementary Information). Since the first DSE (first wave) was a very strong request, it greatly reduced human mobility. Additionally, the mobility reduction was large in the third DSE (fourth wave), in which the requests were strengthened compared to the second DSE to some extent. In comparison, the magnitudes of the responses in the second and fourth DSEs (third and fifth waves), in which the requests were somewhat mitigated, are low. This result differs substantially from the findings of similar studies [25, 26], showing that responses decreased with each DSE. However, our result coincides with that of the sub-analysis in [25] applying spatial structure in the error component.

Our study is the first to demonstrate that responses to information about COVID-19 infection also arise from cross-prefectural travel. These spatial spillovers were remarkable when the public feared and cautioned the new infectious disease and its more infectious variants (in the first and fourth to sixth waves). However, there is no evidence of DSE and vaccination spatial spillovers; this is likely because both are only valid in one’s own prefecture. This lack of spatial effects in NPIs is also observed in another study, which finds that the spatial spillover of NPIs to neighbourhoods has only limited effects on human mobility [8].

We acknowledge that this study possesses the following limitations:

1. The data, which are macro data aggregated by prefecture, do not uncover the diversity of behaviours within a prefecture or for each individual.
2. It does not consider policy measures other than emergency declarations, such as semi-state of emergency COVID-19 measures.
3. Although data on each variant’s infection speed and severity are crucial for the human mobility response, we cannot incorporate this information into our model because of the complexity of the spreading mechanisms and the lack of data on people’s responses to individual information for each variant. Human mobility responses to this information are most likely included in the responses for the number of infections.

Even considering the limitations noted above, our results have several key policy implications. If it becomes necessary to control human mobility during a future pandemic, flexible policies monitoring human mobility responses to information are desirable. One prominent response is that, under repeated waves of similar infection, a larger-than-expected going-out behaviour is expected due to people’s habituation. In addition, the infection rate in other regions, which also affects human mobility, should be considered. Regarding the response to human mobility control measures, people seemed sensitive to the intensity of mobility control policy in Japan. Therefore, fine-tuning policies of mobility control is possible. Implementing flexible human mobility control policies by precisely monitoring human mobility can thus prevent excessive or insufficient mobility control requests. Such a flexible policy could efficiently suppress infection spread and prevent economic activity reduction more than necessary. These implications are useful for evidence-based policymaking during future pandemics.

## Data Availability

The datasets generated during and/or analysed during the current study are available from the author on reasonable request.

## Acknowledgements

The author thanks Takeshi Miura, Dung Luong Anh, Chigusa Okamoto, Yasuharu Ukai, Shigeki Kano, Yuta Kuroda, Hiroshi Uno, Yasutomo Murasawa, and participants in the seminar held at the Osaka Metropolitan University for their helpful comments and suggestions. Any remaining errors are the author’s alone.

## Author contributions

SF conceived the idea for this study and created a project plan, designed the empirical strategy, and acquired and processed the data. SF also performed visualisation and analysis, interpreted findings, and drafted this manuscript.

