## Supplementary Information for "Unravelling habituation for COVID-19-related information: A panel data study in Japan"

Shinya Fukui*

*.

### Appendix A Data description

#### Human mobility

Since human mobility, the dependent variable in the estimation, has day-of-week fluctuations (i.e. each day of the week has its own variation characteristics, such as large variations during weekends), we take the difference from the previous week of the percentage change from the baseline. By taking the week-on-week difference, rather than using the percentage change from the baseline itself, we can capture the effect of new information in the short term, such as a week-on-week change in the number of daily newly infected cases, on people's decisions about whether to go out. An example of a dependent variable is shown in Fig A1.

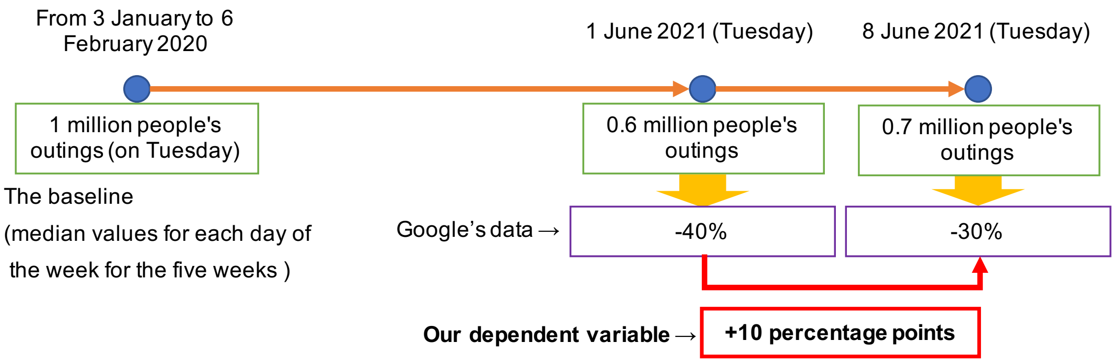


**Fig A1. Example of a dependent variable** The author created fake data for the baseline, 1 June 2021 (the previous week), and 8 June 2021.

#### Infected cases of COVID-19

Data on the daily number of newly infected cases of COVID-19 are obtained from NHK (*NIPPON HOSO KYOKAI*; Japan Broadcasting Corporation) [1]. Given that the number of new cases fluctuates over a vast range, it is better to take the logarithm of the data to mitigate heteroscedasticity. Since the data contain zeroes, we use an inverse hyperbolic sine (IHS) transformation [2, 3, 4, 5], which converts zeros to zeros and behaves similarly to a logarithm. Let $I_{it}$ denote new cases; then, the IHS transformation of $I_{it}$, $I_{it}^*$ becomes

$$I_{it}^* = \ln\left(I_{it} + \sqrt{I_{it}^2 + 1}\right).$$

Additionally, since new cases have day-of-week fluctuations (e.g. fewer PCR tests on weekends), we convert the IHS transformation of new infections to the difference from the same day of the previous week. Hence, the week-on-week difference of daily new infected cases transformed by the IHS, $\Delta I_{it}^{*}$, approximates the growth rate of new cases compared to the previous week.

According to an NHK news article [6], 'A record number of cases—5,773—were confirmed in Tokyo on Friday, 13 August 2021. This is the highest number ever recorded. The number of cases has increased by 1,258 since last Friday, and the rapid spread of infection continues' (translated from the Japanese article by the author). Daily news in Japan primarily covered the number of daily new infections and week-on-week changes in the number of infections. In this way, our approximation of the week-on-week growth rate of new cases in our data for our estimation reasonably illustrates the information received by people judging the severity of the situation based on week-on-week changes in the number of infections.

**The declarations of a state of emergency**

The DSE data are obtained from the Cabinet Secretariat's COVID-19 Information and Resources [7]. The timing of DSEs varied between prefectures. Fig A2 displays DSE periods for each prefecture.

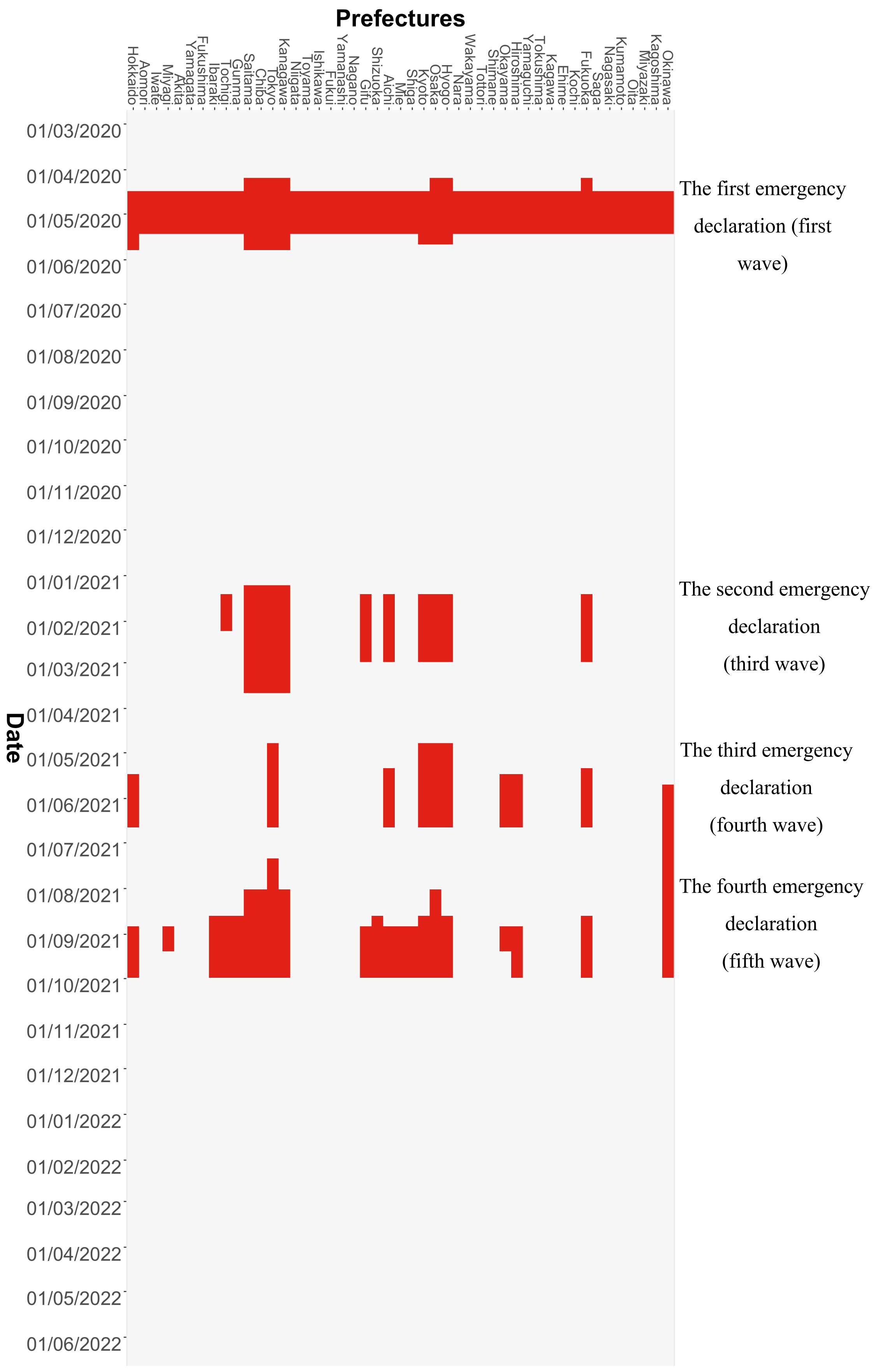
Prefectures
Hokkaido
Aomori
Iwate
Miyagi
Akita
Yamagata
Fukushima
Ibaraki
Tochigi
Gunma
Saitama
Chiba
Tokyo
Kanagawa
Niigata
Toyama
Ishikawa
Fukui
Yamanashi
Nagano
Gifu
Shizuoka
Aichi
Mie
Shiga
Kyoto
Osaka
Hyogo
Nara
Wakayama
Tottori
Shimane
Okayama
Hiroshima
Yamaguchi
Tokushima
Kagawa
Ehime
Kochi
Fukuoka
Saga
Nagasaki
Kumamoto
Oita
Miyazaki
Kagoshima
Okinawa
Date
01/03/2020
01/04/2020
01/05/2020
01/06/2020
01/07/2020
01/08/2020
01/09/2020
01/10/2020
01/11/2020
01/12/2020
01/01/2021
01/02/2021
01/03/2021
01/04/2021
01/05/2021
01/06/2021
01/07/2021
01/08/2021
01/09/2021
01/10/2021
01/11/2021
01/12/2021
01/01/2022
01/02/2022
01/03/2022
01/04/2022
01/05/2022
01/06/2022
The first emergency declaration (first wave)
The second emergency declaration (third wave)
The third emergency declaration (fourth wave)
The fourth emergency declaration (fifth wave)

**Fig A2. DSE periods for each prefecture.** The red colour indicates the period during which the DSE was issued.

### Details of DSE

*NPIs in Japan*

Regarding NPIs in Japan, lockdowns accompanied by legal enforcement have not been implemented. Instead, Japan implemented DSEs that requested that people refrain from leaving their residences except for essential and urgent purposes and that facilities shorten their hours of operation or temporarily close down. As such, the Japanese policies for controlling the spread of COVID-19 are referred to as ‘soft lockdowns’ [8] or ‘voluntary lockdowns’ [4, 5]. This means that even if a DSE is declared, individuals are not legally regulated but are requested to stay at home; accordingly, going out behaviours are voluntary [9]. To account for this, we consider DSEs by the Japanese government to be COVID-19-related information.

*Requests regarding DSEs*

The content of requests regarding DSEs is entrusted to each prefecture and varies by prefecture. This section provides an overview of the details of such requests, focusing on Tokyo and Osaka, Japan’s two representative major cities. Since both are large cities, the requests were almost identical.

In the first DSE (first wave), the closure of facilities and stores was requested in principle for amusement facilities, universities, tutoring schools, educational (school) facilities, exercise facilities, theatres, gathering and exhibition facilities (including museums), and commercial facilities (including department stores). In principle, the government asked that events be held without spectators.

In the second DSE (third wave), the government asked facilities and stores to reduce their operating hours instead of closing entirely. The maximum attendance for events was set at 5,000 people, easing the limitation imposed in the first DSE.

The third DSE (fourth wave), in which the Alpha variant was prevalent, resulted in a new request to close restaurants and stores serving alcoholic beverages and offering karaoke. In addition, requests to close facilities and stores were issued to amusement facilities, exercise facilities, theatres, commercial facilities (including department stores), and museums with footprints of over 1,000 square metres. Additionally, the government asked that events that attract visitors be held without spectators and that college classes be conducted online. Thus, with the emergence of the Alpha variant, the requests were strengthened.

In the fourth DSE (fifth wave), the government asked restaurants and stores serving alcoholic beverages and offering karaoke to close again. Meanwhile, facilities and stores were not requested to close in principle but were asked to shorten their operating hours. Moreover, the maximum attendance for events was again set to 5,000 people. Thus, these requests were less restrictive than those made during the third declaration.

During all the DSEs, restaurants were asked to shorten their operating hours, and people were asked to avoid unnecessarily leaving their homes. The above information regarding the DSEs was taken from the Tokyo Metropolitan Government [10] and the Osaka Prefectural Government [11].

The first DSE was issued to all 47 prefectures. In contrast, the subsequent DSEs were only issued to some prefectures based on the local severity of infections; specifically, the second DSE was issued in 11 prefectures, the third in 10 prefectures, and the fourth in 21 prefectures. Moreover, the length of the DSE periods differed by prefecture. One exception is that, only in Okinawa Prefecture, DSEs were declared for the third and fourth waves without interruption. In all cases, the government asked facilities and stores to close only in the first and third DSEs. In short, the first and third requests were typically strict, and the second and fourth requests were typically lenient; across prefectures.

**Vaccination rates**

The daily data on COVID-19 vaccination of each prefecture are obtained from the COVID-19 Vaccination Status by the Digital Agency [12]. We convert the data into a cumulative format to determine the vaccination rate per million persons. Population data for each prefecture (on 1 October 2020) are obtained from Population Estimates by the Statistics Bureau of Japan [13]. Since the number of vaccinations has day-of-week fluctuations, the data (vaccination rate per million persons) are converted to the week-on-week change.

We have other reasons for utilising week-on-week rather than daily data itself in our estimations. Since vaccinated people have more reassurance that they can avoid infection, we reason that higher vaccination rates are likely to correlate with a higher portion of people going out in each prefecture. In the extreme, each vaccinated individual will shift from the fearful group (unvaccinated group) to the reassured group (vaccinated group). The size of the reassured group will thus increase in step with the promotion of vaccination in the prefecture, and the going-out behaviour of the group will also increase proportionally. Thus, one might think that the better choice would be to use the vaccination rate itself, which ranges from 0–100%.

However, we are using week-on-week differences in retail and recreation behaviour and residential time as the dependent variables in our estimation. Since fear is expected to decrease considerably immediately after vaccination, the growth from the previous week in going-out behaviour is expected to increase only at this point. In other words, individuals who already feel reassured and engage in going-out behaviour right after being vaccinated will essentially not contribute to later growth in going-out behaviour. Therefore, we use the difference in the vaccination rate compared to the previous week (incidentally, when we use the vaccination rate instead, none of the results would have been significant). Theoretically, each vaccinated person shifted from the fearful group to the reassured group and contributed to the growth in going-out behaviour at that point. For example, in the fake data in Fig A1, we can see that the number of people going out from 1 June to 8 June 2021 increased by 0.1 million. If we hypothesise that this increase is entirely due to vaccination, then we can assume that the increase only happens from 1 June to 8 June 2021 and that the 0.1 million newly vaccinated people who joined the reassured group at this time will continue to go out and not contribute to later increases in going-out behaviour.

The other reason we use the difference from the previous week was the multicollinearity between the first and second vaccination doses. Japan's vaccination rate accelerated in 2021 [14]. At the time, due to the Ministry of Health, Labour, and Welfare announcement that two doses of the COVID-19 vaccine can prevent infection [15], most people who received a first dose also received a second dose. Therefore, the first and second vaccination rates almost perfectly correlate (correlation coefficient = 0.99).

**Time series plots**

The time series plots of variables used in the estimation are shown in Fig A3. From the figure, infections and human mobility are moving in opposite directions. Further, the first and second vaccinations proceeded rapidly. Additionally, the timing of the DSE differs by prefecture.

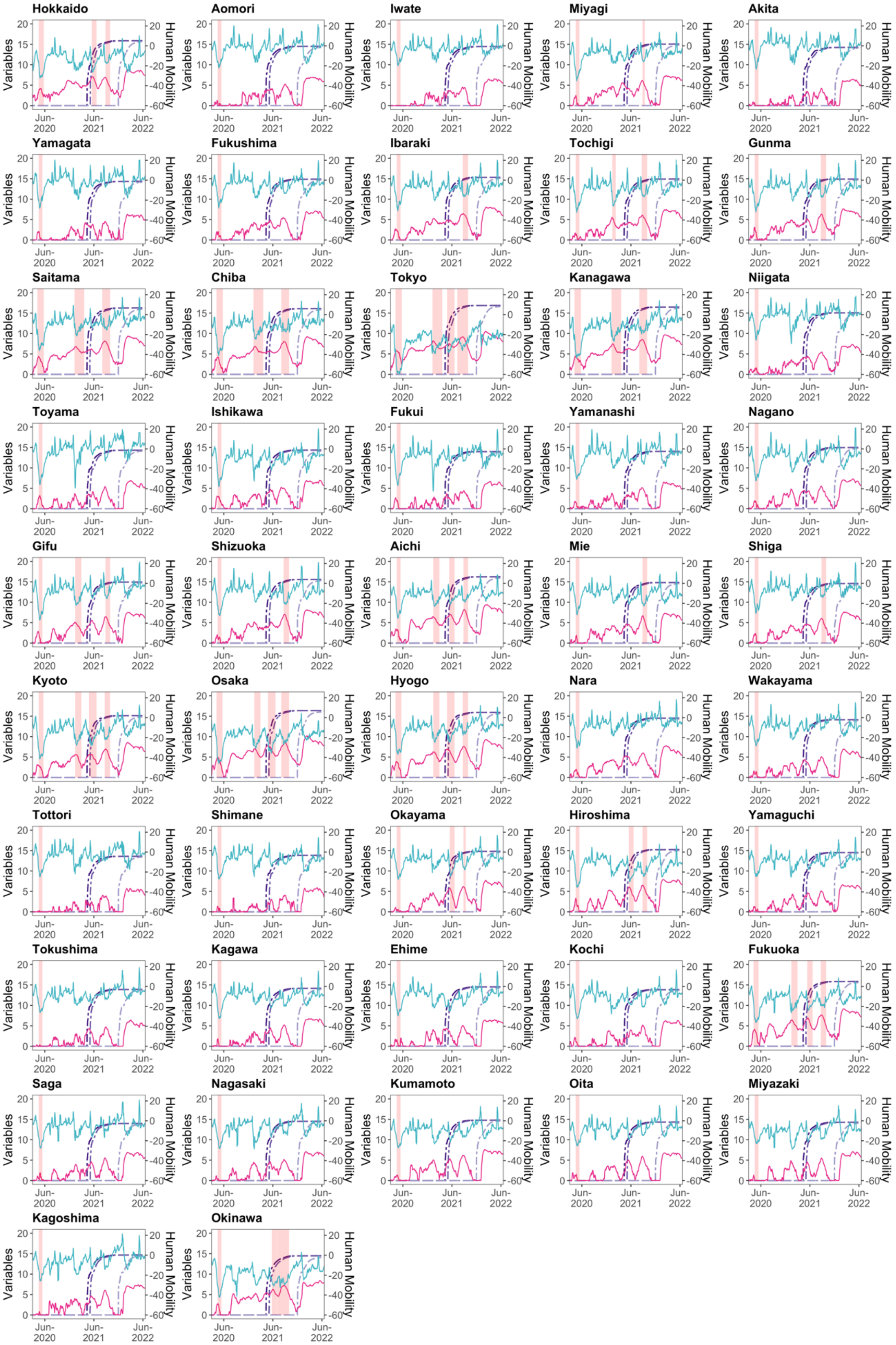
Hokkaido
Aomori
Iwate
Miyagi
Akita
Yamagata
Fukushima
Ibaraki
Tochigi
Gunma
Saitama
Chiba
Tokyo
Kanagawa
Niigata
Toyama
Ishikawa
Fukui
Yamanashi
Nagano
Gifu
Shizuoka
Aichi
Mie
Shiga
Kyoto
Osaka
Hyogo
Nara
Wakayama
Tottori
Shimane
Okayama
Hiroshima
Yamaguchi
Tokushima
Kagawa
Ehime
Kochi
Fukuoka
Saga
Nagasaki
Kumamoto
Oita
Miyazaki
Kagoshima
Okinawa
Variables
Human Mobility
Jun-2020
Jun-2021
Jun-2022

**Fig A3. Time series plots of the variables per prefecture used in the estimation.** On each chart, the blue-green-coloured line (on the right-hand axis) is the 7-day backward moving average using the geometric mean of human mobility in retail and recreation; the red-purple-coloured line is the IHS transformation of the 7-day backward moving average number of infected persons; the purple-coloured double-dashed lines are the IHS transformation of the cumulative number of people vaccinated with 1–3 doses, and the pink-shaded areas are the DSE periods. The data transformation employed here, such as the 7-day backward moving average, is only for visualisation purposes; we use other transformations in our estimation.

### Control variables

Our estimation model employs control variables. Specifically, we exploit temperature (average daily temperature) and precipitation (total daily precipitation) data from the Japan Meteorological Agency [16], which are relevant to human mobility. We select the capital of each prefecture as the geographical location for the daily temperature average and daily precipitation totals (there are only four missing data which we substitute with data from the nearest location). We take the difference from the previous day for the temperature. Additionally, we use the IHS transformation for these two variables.

Day-of-the-week and weekends-and-holidays dummies (abbreviated as holidays-dummies) account for daily fixed effects. The day-of-the-week dummies are Tuesday, Wednesday, Thursday, and Friday. Holidays-dummies take a value of 1 if the day is Saturday or Sunday, a Japanese holiday, Lantan festival (O-bon in Japanese, which took place on 13–16 August for 2020, 2021, and 2022), or New Year's holiday (from 29 December to 3 January, when most businesses and government offices are on vacation); otherwise, they take a value of 0.

As for prefectural fixed effects, two demographics—the population density per square kilometre of inhabitable land area and the percentage of the population over 65 years old—are extracted from the Regional Statistics Database (System of Social and Demographic Statistics) from the Statistics Bureau of Japan [17]. We take logarithms for these two demographics.

### Appendix B Methods

#### Lags in estimation

We take lags for the IHS difference (from the previous week) of new infections, $\Delta I_{it}^*$; its spatially weighted variables, $\boldsymbol{W}_t^* \Delta \boldsymbol{I}_t^*$; the week-on-week increased range of vaccination rate per million persons, $\Delta \boldsymbol{V}_{vt}$; and its spatially weighted variables, $\boldsymbol{W}_t^* \Delta \boldsymbol{V}_{vt}$, in the estimation equation (2) in the main manuscript. Daily lag $p$ from day $t$ is taken from 1 day ($p = 1$) to 7 days ($p = 7$). We also take a lag for the spatial weight matrix, $\boldsymbol{W}_t^*$, which corresponds to lag day $t - p$ of the estimation.

During the pandemic, most people decided whether to go out based on information regarding the infection status announced up to the previous day. The same is true for vaccination $\Delta V_{it}$ , whereby outgoing behaviour is determined by the vaccines administered up to the previous day. Therefore, in our estimation, the maximum lag days is set to 7. By contrast, the lag is not taken for DSE ($E_{it}$), as DSE on the day, rather than the day before, influences outgoing behaviour. Similarly, lags are not taken for the control variables: temperature, precipitation, day-of-the-week dummies, and holidays-dummies, since each is only relevant for human mobility of that day.

Another reason to take the lags for the infected cases is the announcement timing. Daily infected cases are only reported in the evening (around 17:00) or later. For instance, the infected case in Tokyo on 31 August 2021 was announced by NHK at 23:25, Nikkei at 17:00, Bloomberg at 17:19, Jiji Press News at 22:34, TV Asahi at 18:45, and Tokyo Shinbun at 16:56 (based on Google search which accessed on 15 August 2022).

Due to the data used to create the weight matrix, the value is fixed for a given week. Therefore, in our estimation, we use a spatial weight matrix ($\boldsymbol{W}^*$) for the week corresponding to day $t$ in the analysis, which gives $\boldsymbol{W}_t^*$. For example, 15 June 2022 corresponds to the 24th week of 2022. If we take $p = 1$ lag in our estimation, we obtain 14 June 2022, which also corresponds to the 24th week of 2022. Therefore, we choose the spatial weight matrix for the 24th week. Nevertheless, if lag $p = 5$ is taken from 15 June 2022, the observed date would be 10 June 2022, corresponding to the 23rd week of 2022, that is, the spatial weight matrix will be that of the 23rd week. Therefore, the value of the spatial weight matrix does not change by day within the same week.

Estimates are conducted separately for each lag day: meaning that the estimation is performed seven times. A model encompassing all lag orders (distributed lag model) is also estimated to ensure robustness. We employ the polynomial degree 1 Almon lag model to avoid multicollinearity arising from the distributed lag model. Further details on the Almon lag model estimation are provided in Appendix D.

**Constructing a spatial weight matrix in the estimation model**

To construct a spatial weight matrix, $\boldsymbol{W}^*$, we acquire a dataset called Cross-Prefecture Travel Data from Vital Signs of Economy-Regional Economy and Society Analyzing System (V-RESAS) provided by the Cabinet Secretariat and the Cabinet Office, Government of Japan [18]. These data are constructed from Agoop Corporation's Current Population Data, which is based on GPS data obtained with user consent from specific smartphone applications and makes demographic data using day/night population data.

There are two types of data: *movement from other prefectures to the relevant prefecture* and *movement from the relevant prefecture to other prefectures*. In this study, we choose the former. Since the latter is the movement of people in one's own prefecture, reverse causality from the dependent variable (human mobility in one's own prefecture) to the spatial weight matrix (human movement from one's own prefecture to other prefectures) can occur, which creates an endogeneity concern. Furthermore, there are two types of population movement: composition (%) and index. We choose the index because the index allows us to capture the decrease in movement compared to 2019 and the changes in the inter-prefecture movement for each prefecture. The index is based on the average movement across prefectures for all weeks in 2019 as 1. The ISO-8601 week number is employed in the index. According to V-RESAS, the index data is calculated as follows,

$$\text{The index} = \left(\frac{\text{The population that moved from other prefectures to the prefecture during the week in question}}{\text{The average population that moved from other prefectures to the prefecture per week in 2019}}\right).$$

The spatial weight matrix $\boldsymbol{W}$ for the specific week is standardised using Kelejian and Prucha's method [19]:

$$\boldsymbol{W}^* = \boldsymbol{W} \times \frac{1}{\min\left\{\max\limits_{i} \sum_{j}^{M} w_{ij}, \max\limits_{j} \sum_{i}^{N} w_{ij}\right\}}, \text{(A1)}$$

where $w_{ij}$ is an element of the spatial weight matrix with row $i$ (travel from) and column $j$ (travel to), and the diagonal element $w_{ii}$ is 0. This standardisation is conducted for all sample weeks. During the pandemic, the more people travel from prefecture $i$ to their own prefecture $j$, the more the COVID-19 trend in prefecture $i$ is expected to affect human mobility inside $j$ substantially. Two factors can explain this: (1) the higher the interaction of the people between $i$ and $j$, the higher the risk of COVID-19 transmission across prefectural borders; and (2) for commuters $j$ to $i$, the trends in prefecture $i$ are of concern. The larger the element of the spatial weight matrix, $w_{ij}^*$, the higher the number of people moving from prefecture $i$ to $j$.

Using the constructed spatial weight matrix, $\boldsymbol{W}_t^*$, we create the cross-terms, spatially weighted infected cases $\boldsymbol{W}_t^* \times \Delta\boldsymbol{I}_{it}^*$, spatially weighted DSE $\boldsymbol{W}_t^* \times \Delta\boldsymbol{E}_{it}$, and spatially weighted vaccination $\boldsymbol{W}_t^* \times \Delta\boldsymbol{V}_{it}$ to examine the impact of information from the number of infections in other prefectures, the DSE in other prefectures, and the vaccination rate in other prefectures, respectively.

Regarding the impact of NPIs on human mobility, Ilin et al. [20] investigate the spatial spillover effects of NPIs on neighbourhoods. The authors consider certain distances for measuring the effects but not spatial interactions using spatial weights. Another study [21] uses a spatial error panel model to assess the effects of DSE on mobility. However, it does not contain the spatial structure for the explanatory variables, as we do. In addition, in constructing spatial weights, they use a nearest neighbour dummy that captures whether the other prefecture is close or not to a specific prefecture. Our study differs from these studies in that we employ spatial weight matrices constructed by the data related to human travel across prefectures, thus accounting for spatial interactions.

As an example, the elements of the spatial weight matrix for the last week of January 2020 (27 January–2 February 2020) are illustrated in Fig B1. This period occurred just prior to the pandemic when irregular movements due to the New Year celebrations in Japan had already dissipated; as a result, this week is representative of normal inter-prefecture travel. In Fig B1, the dark-red-coloured cells indicate that more people travel between prefectures located in or around large cities such as Tokyo, Aichi, Osaka, and Fukuoka.

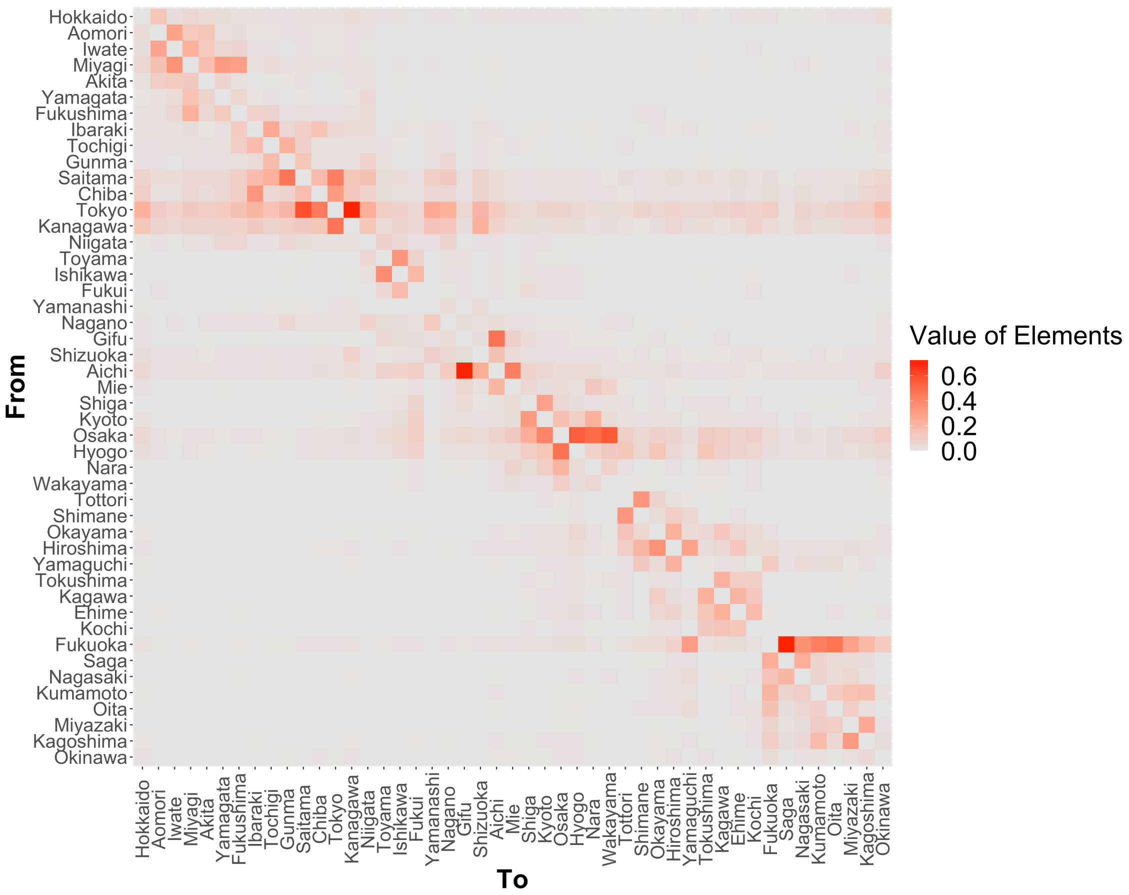


**Fig B1. Spatial weight matrix per prefecture for the last week of January 2020 (27 January–2 February 2020).** Constructed using V-RESAS's Cross-Prefecture Travel data from the Cabinet Secretariat and the Cabinet Office, Government of Japan. Within the figure, the darker the red colour, the greater the number of travellers between prefecture

### Spatially weighted fixed effects in the estimation model

To control for the unobservable spatial spillovers resulting from the travel between each prefectural pair (other than spatially weighted infection, spatially weighted DSE, and spatially weighted vaccination, which are observable), we use the elements of a standardised spatial weight matrix $\boldsymbol{W}^*$, as if the least squares dummy variable (LSDV) estimation. In other words, component $\sum_{j=1}^{M} \delta_j w_{ijt}^*$ represents spatially weighted fixed effects [22, 23], where cross-prefecture travel in each prefecture, $w_{ijt}^*$, varies over cross-sectional (prefectural) dimension $i$ and time dimension $t$ and $\delta_j$ is to be estimated. Unobservable spatial spillovers are related to human travel among prefectures, which are specific to the cross-sectional (prefectural) unit and vary over time.

**Common factors and loadings in the estimation model**

Common factors, denoted by $F_{td}$, are unobservable elements that vary over time and are common for all cross-sectional (prefectural in our case) units. Since each cross-sectional unit (in our case, prefectures) receives a different load from the common factor, $\boldsymbol{\lambda}_d$ describes the difference in loadings. Component $\sum_{d=1}^{D} \boldsymbol{\lambda}_d F_{td_t}$ corresponds to the generalisation of individual-specific and time-specific fixed effects in the panel data analyses; this component can better describe time-varying unobservable elements with different loadings through cross-sectional units than ordinary two-way fixed effects.

In fact, there are unobservable factors affecting human mobility that should be taken into account. For example, suppose a new variant of COVID-19 emerged in a country other than Japan. In that case, it is probable that people in urban areas such as Tokyo and Osaka, which have international airports and large population concentrations, would be more cautious of the outbreak than people in rural areas. Thus, when vigilant of the severity of a new variant infection, people in urban and rural areas will exercise different levels of caution in responding to such information.

Another example of an unobservable factor is how government policies are transmitted. For instance, even if there is an announcement by the Japanese government regarding vaccinations, each prefecture has a different system for promoting vaccinations, so residents in each prefecture (or, more specifically, each municipality, which is the main body promoting vaccinations) will receive the announcement differently. In addition, people may welcome the rapid vaccination programme announcement more in prefectures with higher infection rates.

In such cases, these pieces of information are either unobservable or difficult to incorporate into the model. Additionally, such information affects all prefectures simultaneously; however, the level of sensitivity differs by prefecture. Therefore, the interactive effects model is a better method for controlling these unobservable factors. In the first example, common factors $F_{td}$ capture the risk of epidemics of the new variants, while the loadings $\boldsymbol{\lambda}_d$ capture differences between prefectures in vigilance against the new variants.

A Hausman-type specification test proposed by Bai [24] is used to determine whether it is appropriate to use the factors or classical two-way fixed effect; the results of all tests support the factor type. Dimension $d$ of factors is chosen by consistent estimation (Bai and Ng, [25]), which also considers the underestimation of the true variance. Our results show that dimension $d = 7$. For the variance-covariance matrix, we use heteroscedasticity and autocorrelation consistent estimators, proposed by Bai [24]. The estimation of the interactive effects model is conducted using the 'phtt' R library with circumstances of R 4.0.5.

### Appendix C Duration of the COVID-19 infection wave for each prefecture

As the government made no official announcements regarding the beginning and end of COVID-19 infection waves, we independently determine the COVID-19 wave duration of each prefecture, as shown in Fig C1.

The duration of the COVID-19 infection wave in each prefecture is determined using the 7-day backward moving average of new COVID-19 cases in each prefecture from 22 February 2020 to 15 August 2022. The first wave of infections began on 22 February 2020 (due to data availability). The endpoint of each wave in each prefecture is the day with the lowest number of infections in that wave (in some cases, there may be several days in a row with the lowest number of infections; however, we assume that the subsequent wave begins the day after the first record low). In addition, we assume the wave lasts from when the DSE is issued until it is lifted (the wave does not switch during the DSE, with the exception of Okinawa Prefecture, where the DSE was issued in succession in the third and fourth waves).

In the first wave, in some cases, some prefectures repeatedly moved back and forth between having no infections and having infections. In such cases, it is difficult to determine the lowest infection; therefore, the wave's endpoint is specified so that the wave does not deviate largely from those in other prefectures. In this study, it is necessary to identify the peak of each wave of infections to analyse each wave's increasing and decreasing phases separately. The peak is defined as the day with the highest 7-day backward moving average of new cases in the wave (however, the peak could not be observed only for the first wave in Iwate Prefecture; accordingly, the average of the peaks of all prefectures' waves serves as a proxy). As a result, six waves have been identified. The final point is specified as 20 June 2022, when the wave for all the prefectures (i.e. the entirety of Japan) hit a new low.

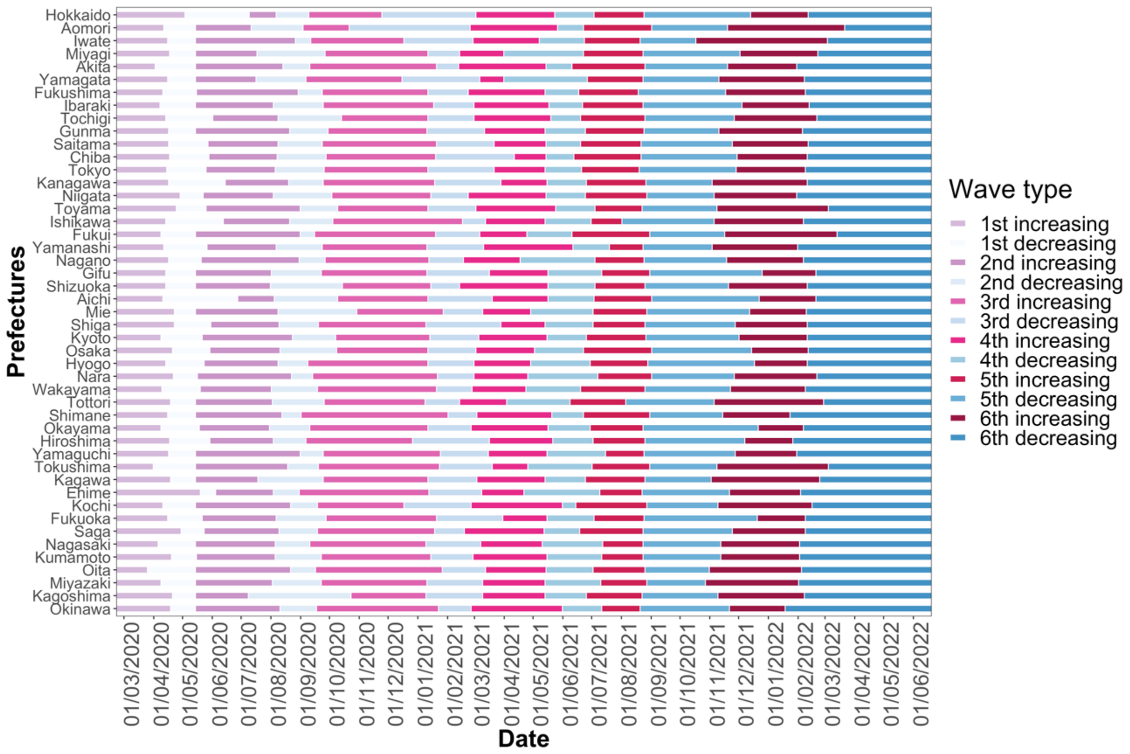


**Fig C1. COVID-19 wave durations in each prefecture.** The data are from 22 February 2020 to 20 June 2022. There are six waves in total, generated by taking a 7-day backward moving average of the number of daily new infections of COVID-19 from NHK.

### Appendix D Performing the estimation through the Almon lag model

When considering the Almon lag model with lags from 1 to 7,

$$y_{it} = a + \sum_{p=1}^{7} b_p x_{it-p} + e_{it}, \text{(A2)}$$

where $i$ is the cross-sectional index, $t$ is the time index, and $p$ is the lag taken for the time dimension $t$. The expression of the polynomial degree 1 Almon lag model is

$$y_{it} = a + \theta_0 \sum_{p=1}^{7} x_{it-p} + \theta_1 \sum_{k=1}^{7} k x_{it-p}\, , \text{(A3)}$$

The estimated coefficients can be retrieved from

$$\hat{b}_p = \hat{\theta}_0 + \hat{\theta}_1 p. \text{(A4)}$$

To find the standard errors of the estimated coefficients, we use the following equation:

$$s.e.(\hat{b}_p) = \sqrt{Var[\hat{b}_p]} = \sqrt{Var[\hat{\theta}_0 + \hat{\theta}_1 p]}$$

$$= \sqrt{\sum_{q=0}^{2} p^{2q} Var[\hat{\theta}_q] + 2 \sum_{q<r} p^{(q+r)} Cov[\hat{\theta}_q, \hat{\theta}_r]}, \text{(A5)}$$

where $q$ and $r$ are the order of $\theta$, that is, the degree of the Almon lag, and $q \neq r$. For a detailed explanation of the Almon lag model, see, for instance, Chapter 17 in [26].

### Appendix E Results of the residential time response

The *residential* category indicates the change in the time spent at home. According to Google [27], given that people are already spending a large portion of their day at their residences, the change in time spent at home is not substantial (even on workdays) compared to the human mobility response. As per Fig E1, the response is minimal; however, the trends are clear.

*E.1 Infected cases in the increasing phase*

A 1% week-on-week increase in the number of infected cases during the first wave is associated with at most a 0.26 pp (lag: 3, s.e. = 0.03) week-on-week increase in the percentage change of the residential time from the baseline (Fig E1a). In the second wave, the confidence interval is narrower, and the magnitude of the increase in the residential time is lower than in the first wave, with a maximum of 0.11 pp (lag: 2, s.e. = 0.02). Similar to the case of retail and recreation, the impact in the first and second waves decreased each day (from 3-day to 7-day lags in the first wave and from 2-day to 7-day lags in the second wave).

In the third and fourth waves, the effect is even more negligible, with a maximum increase of 0.09 pp in the third wave (lag: 3, s.e. =0.02) and a maximum increase of 0.09 pp in the fourth wave (lag: 7, s.e. = 0.04). The fourth wave has a slightly increasing trend as the lag periods are longer. For all four waves, all of the results are significant at a 5% significance level. While for the fifth wave, when the Delta variant was prevalent, the impact increase from lag 1 to lag 4, with a maximum increase of 0.14 pp (lag: 4, s.e. = 0.04). Similarly, for the sixth wave, when the Omicron variant was prevalent, an increasing trend is observed as the lag periods become longer, with a maximum increase of 0.10 pp (lag: 7, s.e. = 0.03).

A habituation trend is also observed for stay-at-home behaviour from the first to the third wave. However, due to the prevalence of the new variants from the fourth wave, the habituation trend halted, and the fourth to the sixth waves show an increasing tendency each lag day, as in the retail and recreation case.

*E.2 Infected cases in the decreasing phase*

In the decreasing phase (Fig E1b), when the estimates are positive and significant, means the residential time decreases as the infected cases drop. In the first wave, residential time is reduced by 0.14 pp at max (lag: 4, s.e. = 0.02). However, the impact is smaller than in the increasing phase. Similar to the retail and recreation case, behaviours did not entirely return to normal, possibly due to fear of COVID-19. In the second wave, the impact is even smaller, with a maximum reduction of 0.08 pp (lag: 2, s.e. = 0.02). At the same time, the third wave is insignificant except for lag 3; furthermore, the impact of lag 3 is negligible. Conversely, in the fourth wave, the magnitude of the estimates increases slightly. Notably, the same tendency (response rose in the fourth wave) is observed in the retail and recreation case, with a maximum reduction of 0.12 pp (lag: 2, s.e. = 0.01). Again, the fifth wave is insignificant for all lag days, consistent with the retail and recreation case. The sixth wave, as in the case of retail and recreation, has a larger impact, with a 0.17 pp reduction (lag: 3, s.e. = 0.05).

*E.3 Spatially weighted infected cases*

For spatially weighted infected cases (Fig E1c), as for spatially weighted infected cases in retail and recreation, the impact of the first wave is large. The maximum impact is 0.46 pp (lag: 6, s.e. = 0.07) on residential time (i.e. residential time increases as the infected case increases in other prefectures and vice versa). In the second wave, the impact drops, with a maximum of 0.11 pp (lag: 2, s.e. = 0.03). In the third wave, all lags are insignificant at a 5% level. In the fourth wave, the estimates are slightly higher, with a maximum of 0.19 pp (lag: 5, s.e. = 0.07). Meanwhile, in the fifth wave, all lags are insignificant except lags 3, 5 and 6; these three lags all have negligible estimates. Finally, the sixth wave, when the Omicron variant spread, shows the influence of infection information from other prefectures—while all lags are significant, the effect is relatively weak.

*E.4 DSE and spatially weighted DSE*

Although the estimates for the DSEs are positive, only the third wave is significant at a 5% level (estimated coefficient = 0.21, s.e. = 0.03) (Fig E1d). DSEs have relatively small impacts on residential time compared to retail and recreation. The spatially weighted DSE is positive and significant for all waves, but the impact is weak (Fig E1e).

*E.5 Vaccination and spatially weighted vaccination*

An increase in vaccination rates within a prefecture negatively affects stay-at-home behaviour (i.e. people become more willing to go out). The results show that only lags 5 through 7 for the second vaccine dose are significant (Fig E1f). Although the magnitude is relatively small, similar to retail and recreation, the second vaccine dose effectively changed human behaviours. While spatially weighted vaccination has negligible effects (Fig E1g).

*E.6 Control variables*

Of the control variables (Fig E1h), only precipitation is positively significant; the more precipitation, the more likely people are to stay at home.

*E.7 Results for the Almon lag model*

The estimated results utilising the Almon lag model for robustness are displayed in Fig E2. The results here generally support the results shown in Fig E1.

*E.8 Comparison between retail and recreation and residential time*

Overall, Fig 3 in the main manuscript and Fig E1 show that, although the magnitude differs, retail and recreation, and residential human behaviours are quite the opposite impacted by COVID-19-related information during each wave. From this, the results for the residential time support our main results for the retail and recreation responses.

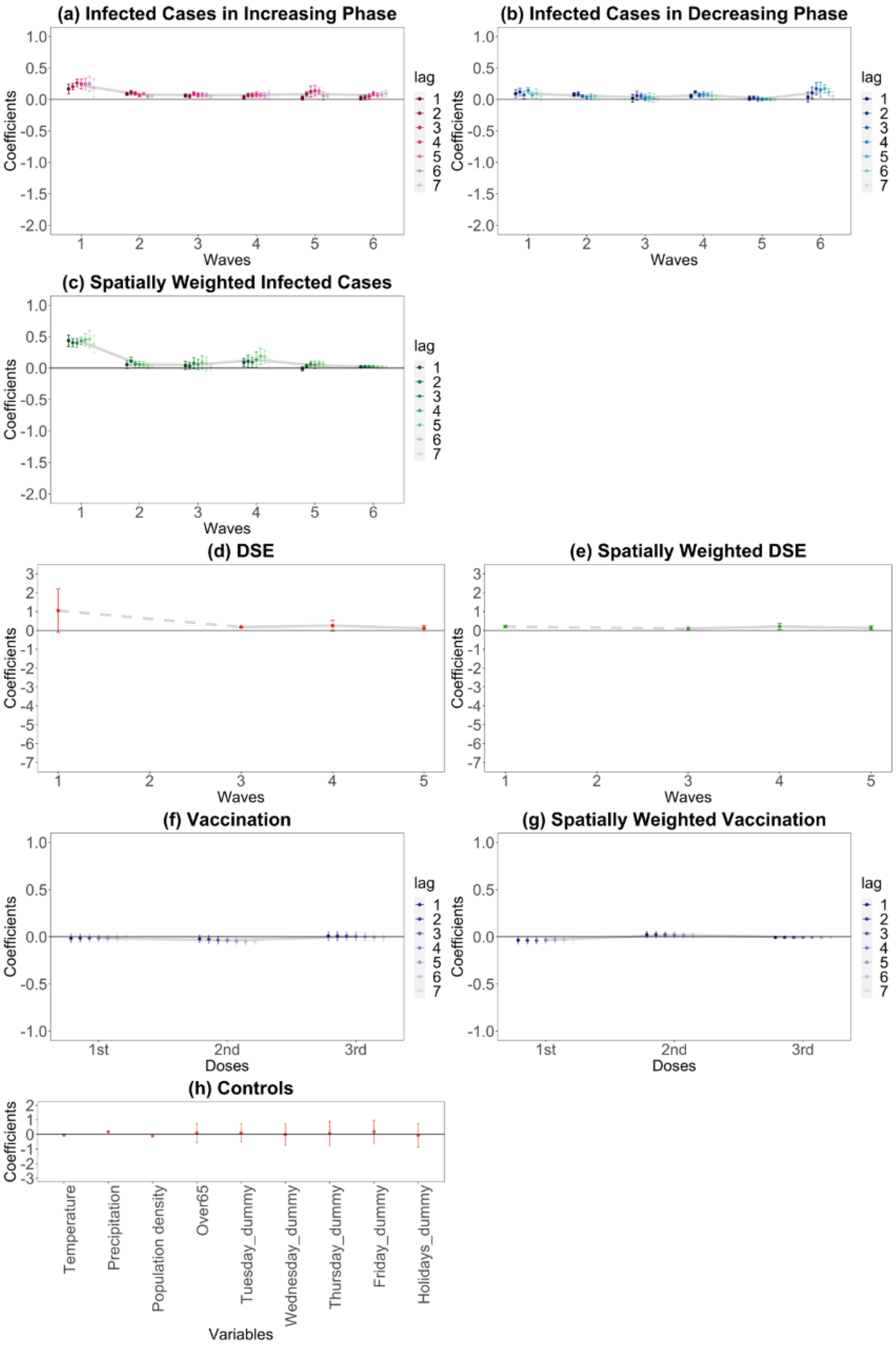
(a) Infected Cases in Increasing Phase
(b) Infected Cases in Decreasing Phase
(c) Spatially Weighted Infected Cases
(d) DSE
(e) Spatially Weighted DSE
(f) Vaccination
(g) Spatially Weighted Vaccination
(h) Controls
Coefficients
Waves
Doses
Variables
lag
1st
2nd
3rd
Temperature
Precipitation
Population density
Over65
Tuesday_dummy
Wednesday_dummy
Thursday_dummy
Friday_dummy
Holidays_dummy

**Fig E1. Residential time responses to COVID-19-related information.** On each chart, the points are estimated coefficients, and the bars indicate upper and lower 95% confidence intervals. The grey line traces the average coefficients of each lag day. There are six infection waves, but DSEs were only issued for the first, third, fourth, and fifth waves. We take a daily lag from 1 to 7 days for infected cases in the increasing phase, infected cases in the decreasing phase, spatially weighted infected cases, vaccination, and spatially weighted vaccination. We conduct the regression analysis seven times, from lags 1 to 7. We do not take a daily lag for the DSE, spatially weighted DSE, and controls; these estimates are from the lag-1 regression.

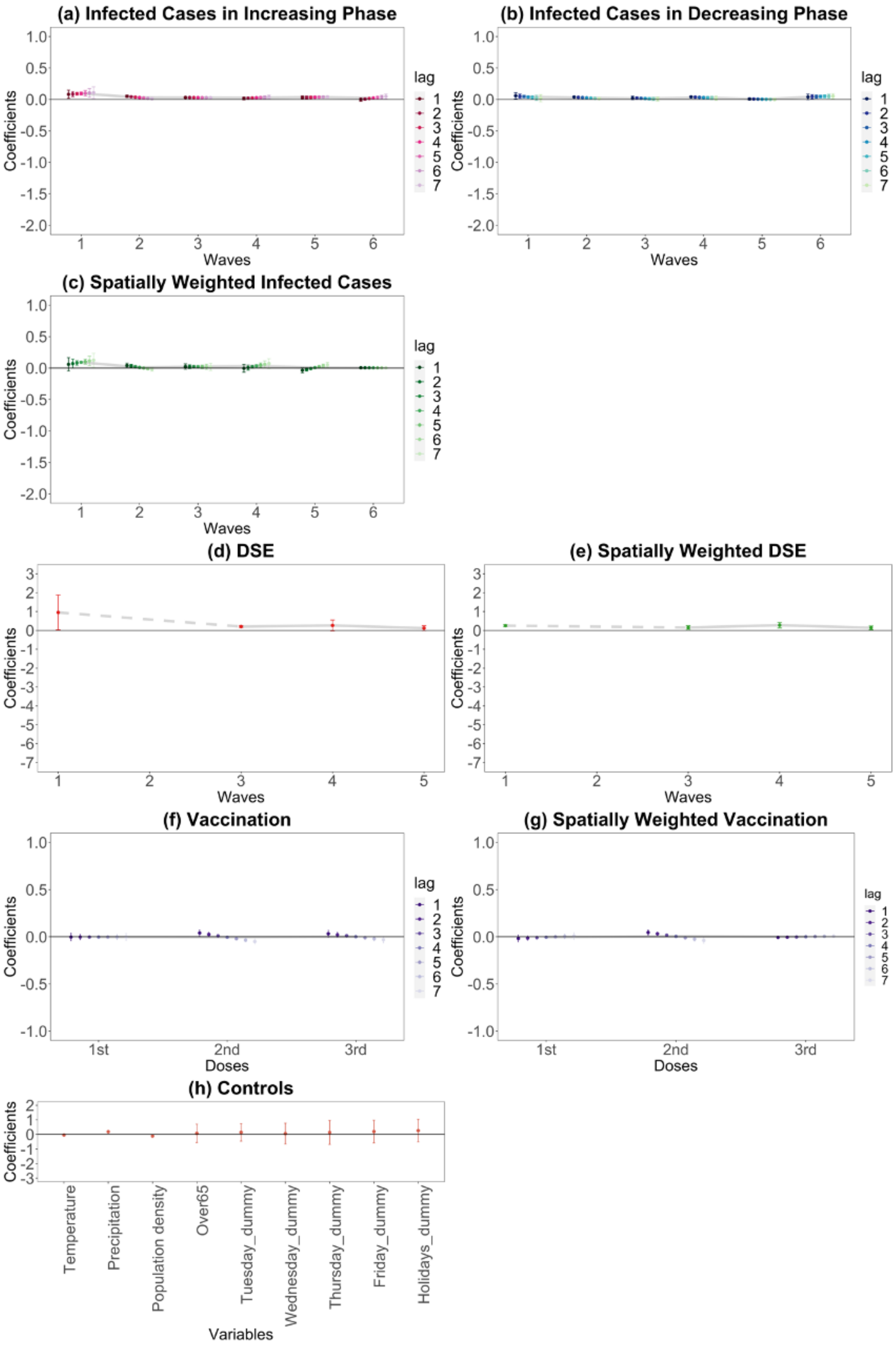
(a) Infected Cases in Increasing Phase
(b) Infected Cases in Decreasing Phase
(c) Spatially Weighted Infected Cases
(d) DSE
(e) Spatially Weighted DSE
(f) Vaccination
(g) Spatially Weighted Vaccination
(h) Controls
Coefficients
Waves
Doses
Variables
lag
Temperature
Precipitation
Population density
Over65
Tuesday_dummy
Wednesday_dummy
Thursday_dummy
Friday_dummy
Holidays_dummy

**Fig E2. Residential time responses to COVID-19-related information using the Almon lag model.** On each chart, the points are estimated coefficients, and the bars indicate upper and lower 95% confidence intervals. The grey line traces the average coefficients of each lag day. There are six infection waves, but DSEs were only issued for the first, third, fourth, and fifth waves. We take a daily lag from 1 to 7 days for infected cases in the increasing phase, infected cases in the decreasing phase, spatially weighted infected cases, vaccination, and spatially weighted vaccination. We do not take a daily lag for the DSE, spatially weighted DSE, and controls.
